# Study of Non-Pharmacological Interventions on COVID-19 Spread

**DOI:** 10.1101/2020.05.10.20096974

**Authors:** Avaneesh Singh, Saroj Kumar Chandra, Manish Kumar Bajpai

## Abstract

COVID-19 disease has emerged as one of the life threatening threat to the society. It is caused by a novel beta coronavirus. It began as unidentified pneumonia of unknown etiology in Wuhan City, Hubei province in China emerged in December 2019. No vaccine has been produced till now. Mathematical models are used to study impact of different measures used to decrease pandemic. Mathematical models have been designed to estimate the numbers of spreaders in different scenarios in the present manuscript. In the present manuscript, three different mathematical models have been proposed with different scenarios such as screening, quarantine and NPIs for estimating number of virus spreaders. The analysis shows that the numbers of COVID-19 patients will be more without screening the peoples coming from other countries. Since, every people suffering with COVID-19 disease are spreaders. The screening and quarantine with NPIs have been implemented to study their impact on the spreaders. It has been found that NPI measures are able to reduce number of spreaders. The NPI measures reduces the growth of the spread function and providing decision makers more time to prepare with in dealing of the disease.

## 1 Introduction

First Corona case has been reported in Wuhan city of Hubei Province in south China on December 31, 2019 as unidentified pneumonia [1, 2]. On 7 January 2020, Chinese government and the World Health Organization (WHO) have identified virus as a novel coronavirus (2019-nCoV) which belongs to the same virus family of the Severe Acute Respiratory Syndrome (SARS) that out broke also in South China in 2002-2003 [3]. Tyrell and Bynoe [4] first describe Coronavirus in 1966. It has four family members namely alpha, beta, gamma and delta. While alpha and b eta coronaviruses apparently originate from bats, gamma and delta viruses originate from pigs and birds. Among these beta coronaviruses may cause severe disease and fatalities, whereas alpha-coronavirus cause asymptomatic or symptomatic infections. Severe Acute Respiratory Syndrome corona virus 2 (SARSCOVID-2) belongs to beta-coronaviruses and is closely related to the SARS-COVID-2 virus. SARS-COVID-2 is 96% identical at the whole-genome level to a bat coronavirus. The most common symptoms of COVID-19 are fever, tiredness, and dry cough [5]. Some patients may have aches and pains, nasal congestion, runny nose, sore throat or diarrhea. Around 1 out of every 6 people who gets COVID-19 becomes seriously ill and develops difficulty breathing [6]. People can be infected of COVID-19 disease from others who have the virus. The disease can spread from person to person through small droplets from the nose or mouth which are spread when a person with COVID-19 coughs or exhales. People can also catch COVID-19 if they breathe in droplets from a person with COVID-19 who coughs out or exhales droplets. There is an increasing body of evidence to suggest that human-to-human transmission may be occurring during the asymptomatic incubation period of COVID-19, which has been estimated to be between 2 to 10 days [7]. On 30th January 2020, the WHO declared the Chinese outbreak of COVID-19 as pandemic [5]. All major counties have suspended all the international flights to reduce international traveler to the country for preventing COVID-19 diseases [6]. At present, no effective antiviral treatment or vaccine is available for COVID-19. The countries have taken strong measures to prohibit the virus’s transmission, such as warning citizens from going outdoors, suspending the public transport between some big cities, and even taking quarantine for the main infected city. These unprecedented measures were expected to effectively stop the virus transmission and buy necessary time to deploy medical resources to the affected area [7]. The rapid spread of the COVID-19 may be due to multiple causes. One cause is the lacking of information transparency at the early stage of the epidemic outbreak. Releasing the epidemic information in a timely and accurate way is extremely important for the anti-epidemic response of the public. The authentic and transparent information could have prohibited the spread of the COVID-19 at the early stage. The other cause is the lacking of scientific diagnostic criterion for the COVID-19. Rapid developing exact testing techniques for a novel virus is very difficult. In fact, the symptoms of the COVID-19 are highly similar to those of flu. This aggravated the hardship of diagnosis. The last but not the least, the lacking of an epidemic warning and prediction system lost the opportunity to prohibit the epidemic spread at the initial stage.

Mathematical modelling has gained more attention and awareness in epidemiology and the medical sciences [8, 9, 10, 11]. It has been used for cancer detection, segmentation and classification [12, 13, 14, 15].These models are useful in the cases where disease dynamics are not unclear. It estimates the number of cases in worst and best-case scenarios. It is also helpful in estimating effect of preventive measures adopted against novel viruses such as COVID-19. One family of these models is the dynamical epidemic model called Susceptible Infected-Removed (SIR) model. The SIR model originated from the study of the plague almost one hundred years ago. Tremendous advance has been achieved in dynamical epidemic model since mid-20th century. In recent decades, some realistic factors influencing the epidemic transmission were included in the classic SIR model, such as the model considering the incubation stage, the SEIRS model considering the population age and the population exposed to epidemic, and the SIS model including birth and death of the susceptible. Some dynamical models were also designed for specific epidemic. In the present manuscript, a novel mathematical model is being presented for estimating novel corona virus patients.

The proposed model also considers Non-Pharmacological interventions (NPIs) such as Social distancing, closure of schools, universities and offices, avoidance of mass gatherings, community-wide containment etc [16, 17]. In addition, the proposed model has been analyzed with the general population, contacts of cases (Contact tracing, Surveillance, quarantine) and the cases themselves (Early Reporting, Isolation). The present work is organized as follows. The mathematical model for COVID-19 spread estimation is presented in section 2. Section 3 discusses data set used for validating proposed mathematical models. Results ad discussions are presented in section 4.

## 2 Proposed Methodology

In the present manuscript, three mathematical models are being presented for estimation of peoples with COVID-19 disease.

1. Mathematical model without screening, quarantine and lockdown
2. Mathematical model for spread with screening and quarantine
3. Mathematical model for spread with screening, quarantine and lockdown

### 2.1 Mathematical model without screening, quarantine and lockdown

The mathematical model for estimating COVID**-**19 patients without screening, quarantine and lockdown is being presented in this section. In this model, all the peoples who are suffering with COVID**-**19 diseases are spreaders. The spreaders are roaming freely around the city and able to spread corona virus to large number of peoples. Hence, the number of estimated peoples with COVID-19 disease will be more. The proposed mathematical model can be defined as:

(case if *m <* 14)

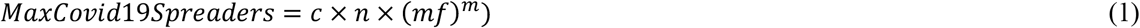

(case if *m >* 14)

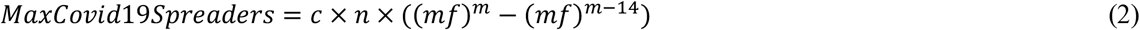

Here, *c* is constant and *n* is initial spreaders. *mf* is multiplying factor of the disease and *m* is number of days.

### 2.2 Mathematical model with spread with screening and quarantine

The mathematical model for estimating COVID-19 patients with screening and quarantine is being presented in this section. In this model, not all the peoples who suffering with COVID-19 diseases are spreaders. Initial screening of new comers is being done for testing COVID-19 diseases. The proposed mathematical model is presented in the Fig. 1. In the Fig. 1, at the top level COVID-19 screening is being done. At this time, peoples are being tested for COVID-19 diseases. Hence, the peoples are divided into two groups namely the peoples with COVID-19 disease and the peoples without COVID-19 disease. The peoples with COVID-19 are quarantined so that they cannot infect other peoples. It is also possible that the COVID-19 symptoms may appear further after initial screening. Hence, these peoples are further divided into two groups namely infectious and non-infectious. The infectious peoples are categorized into three categories based on symptoms such as the peoples having symptoms, the peoples having mild symptoms and the peoples not having any kind of symptoms. The peoples having symptoms are suggested to be self-quarantined. The self-quarantined peoples may be cured or died. The peoples who are not self-quarantined may die or having sufficient immunity to face COVID-19 and virus died after completing period. The peoples having mild symptoms are suggested to be self-quarantined. If they follow quarantine; they may have symptoms which follow the process described previously. The people may die or virus may die after completing virus cycle. The COVID-19 symptoms may be developed further. In that case, it is being classified into the peoples with developed symptoms, the peoples with mild and the peoples without any symptoms. The peoples whose symptoms are not identified may die or survive after completion of virus cycle. These peoples are spreaders and spread corona virus to large number of peoples before they die. Hence, the number of estimated peoples with COVID-19 disease will be more as compared to model presented in the section 1. The proposed mathematical model can be defined as:

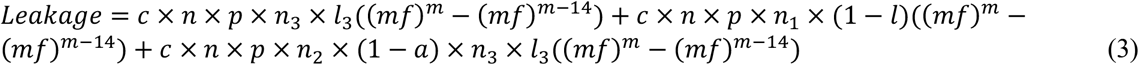

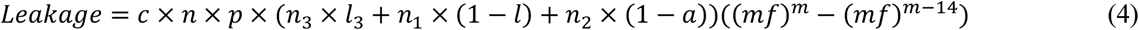

**Figure 1:**
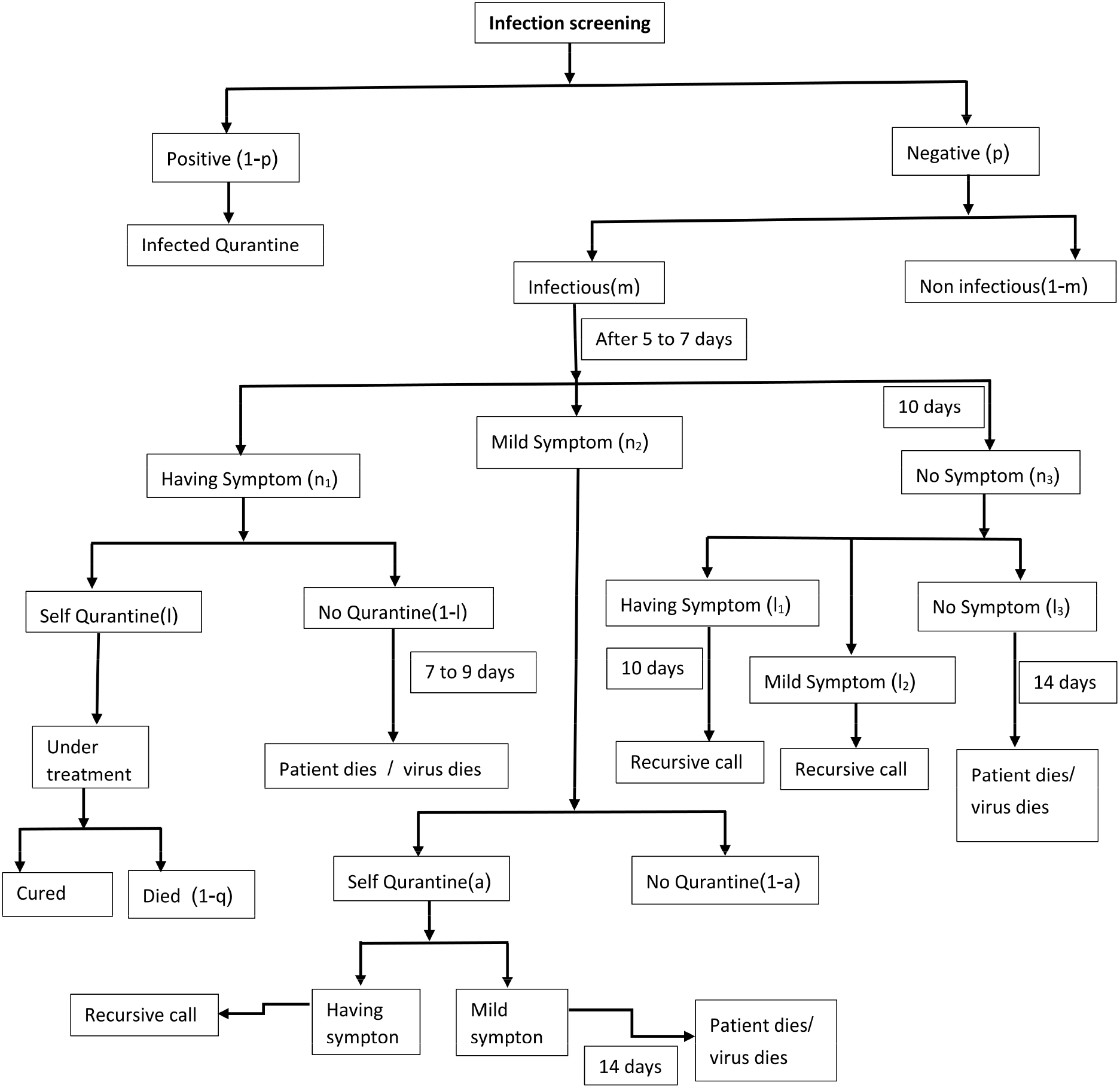
Mathematical Model for CORONA Patient Estimation for spread with screening and quarantine

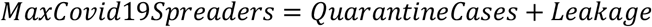

Here, *n*, *m* and *mf* have the same meaning as in model 1. *p* is the fraction of people tested negative in earlier screening. *m*_1_ is the fraction of people who are infectious. *n*_1_, *n*_2_, *n*_3_ are the fraction of people having symptoms, mild symptoms and no symptoms respectively in such a way that sum of *n*_1_, *n*_2_, and *n*_3_ is one. *l* is the fraction of people who have followed self quarantine who have symptoms. *a* is the fraction of people who have followed self quarantine having mild symptoms. *l*_1_, *l*_2_, and *l*_3_ are the portion of people initially having no symptoms but after due time they have been emerged as having symptoms, mild symptoms and no symptoms, respectively.

### 2.3 Mathematical model with spread with screening, quarantine with NPIs

The mathematical model for estimating COVID-19 patients with screening, quarantine with complete NPI measures. Non-Pharmacological Intervention (NPI) is intended to reduce transmission of disease to healthy peoples. The NPI includes distancing measures like no handshaking, working from home, closing of educational institutions, cancellation of mass gatherings, partial or total closure of malls/ markets closure of public transportation and voluntary civic shutdown, “Janata curfew” etc. In this model, not all the peoples who suffering with COVID-19 diseases are spreaders. The proposed mathematical model is same as presented in the Fig. 1. However, in this model, NPI measures are being implemented to reduce number of spreaders. The estimated number of peoples with COVID-19 diseases will be less than that the model presented in the section 2.1 and 2.2. This model focuses on reducing the leakage term presented in model 2. NPI measures reduces the number of persons, which can come in contact with the spreaders and hence reduces the leakage.

## 3 Validation Data Set

Table 1 tabulates COVID-19 data for India [18]. It has various fields such as date, time, active cases, total cured cases, total death cases, total cases, new case, daily death cases and daily cured cases. Here, date and time field describe the COVID-19 cases at particular date and time. Active cases and cured cases are the number of peoples with COVID-19 disease and cured COVID-19 disease cases respectively. Total death field describe number of peoples died due to COVID-19 disease. Total cases field is sum of active cases, death cases and cured cases. Total death field describe number of deaths due to COVID-19 disease. The spreaders are effected by external factors such as suspended international flights, curfew, quarantine and gatherings and lockdown.

**Table 1:**
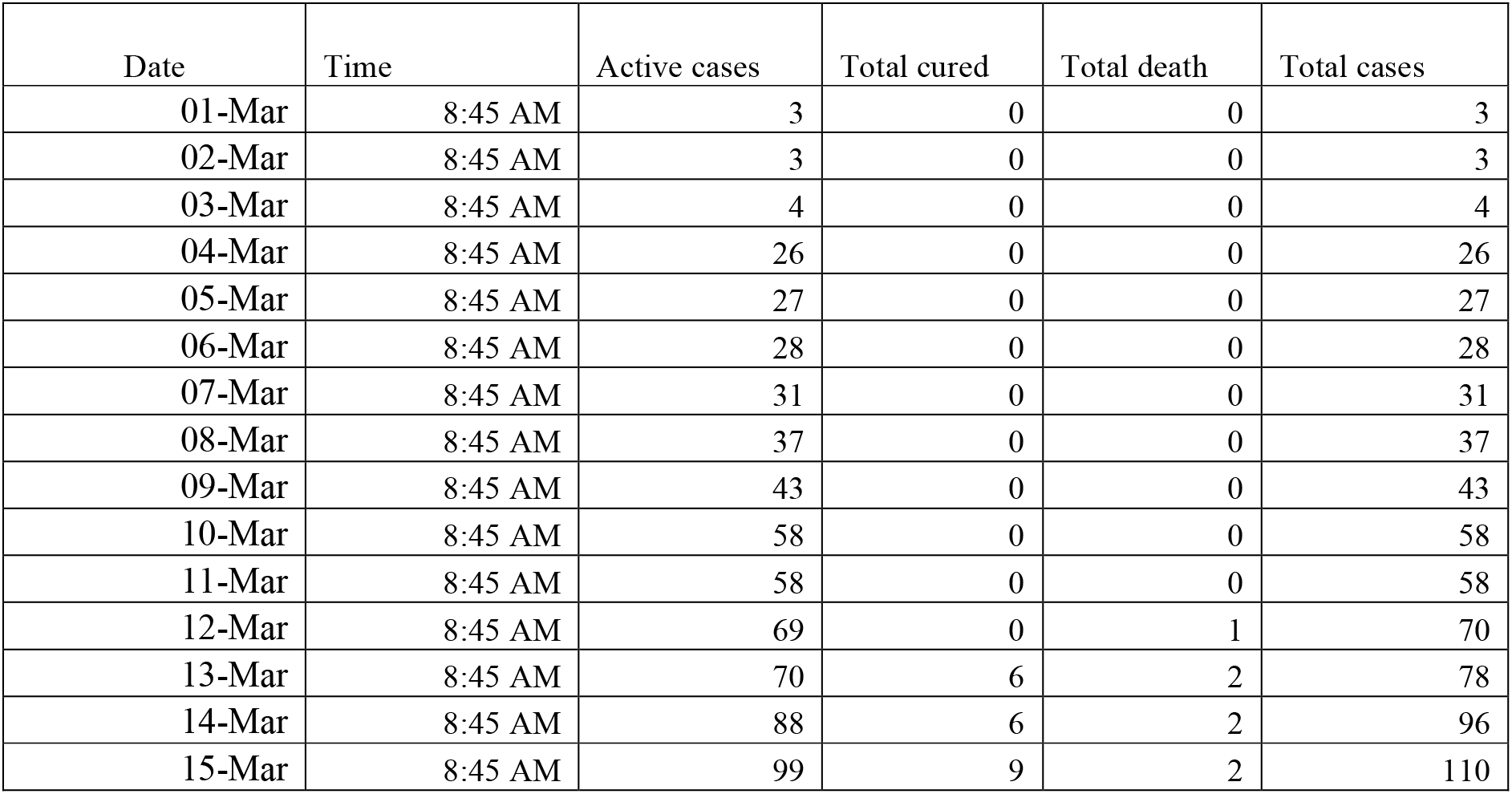
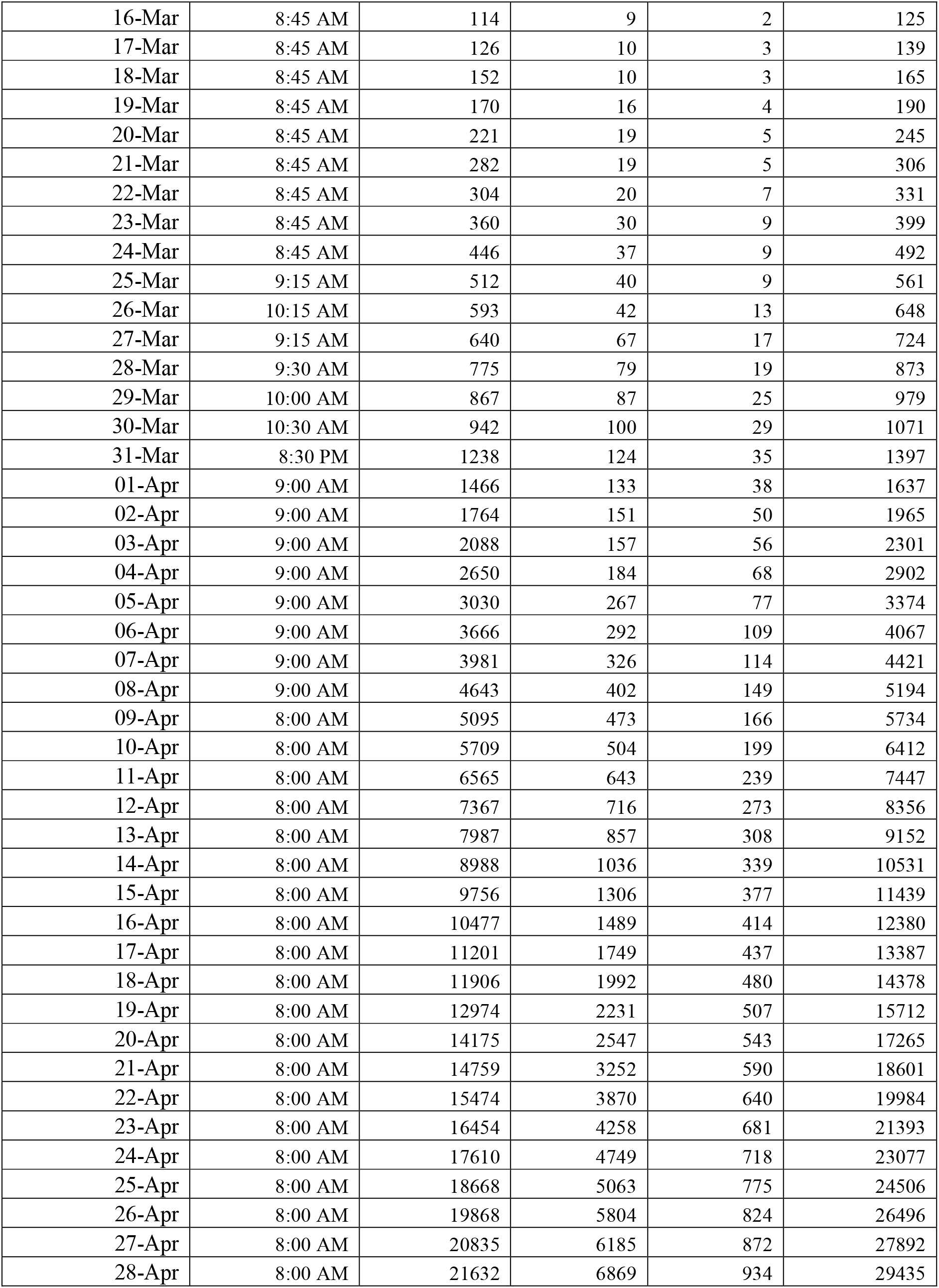
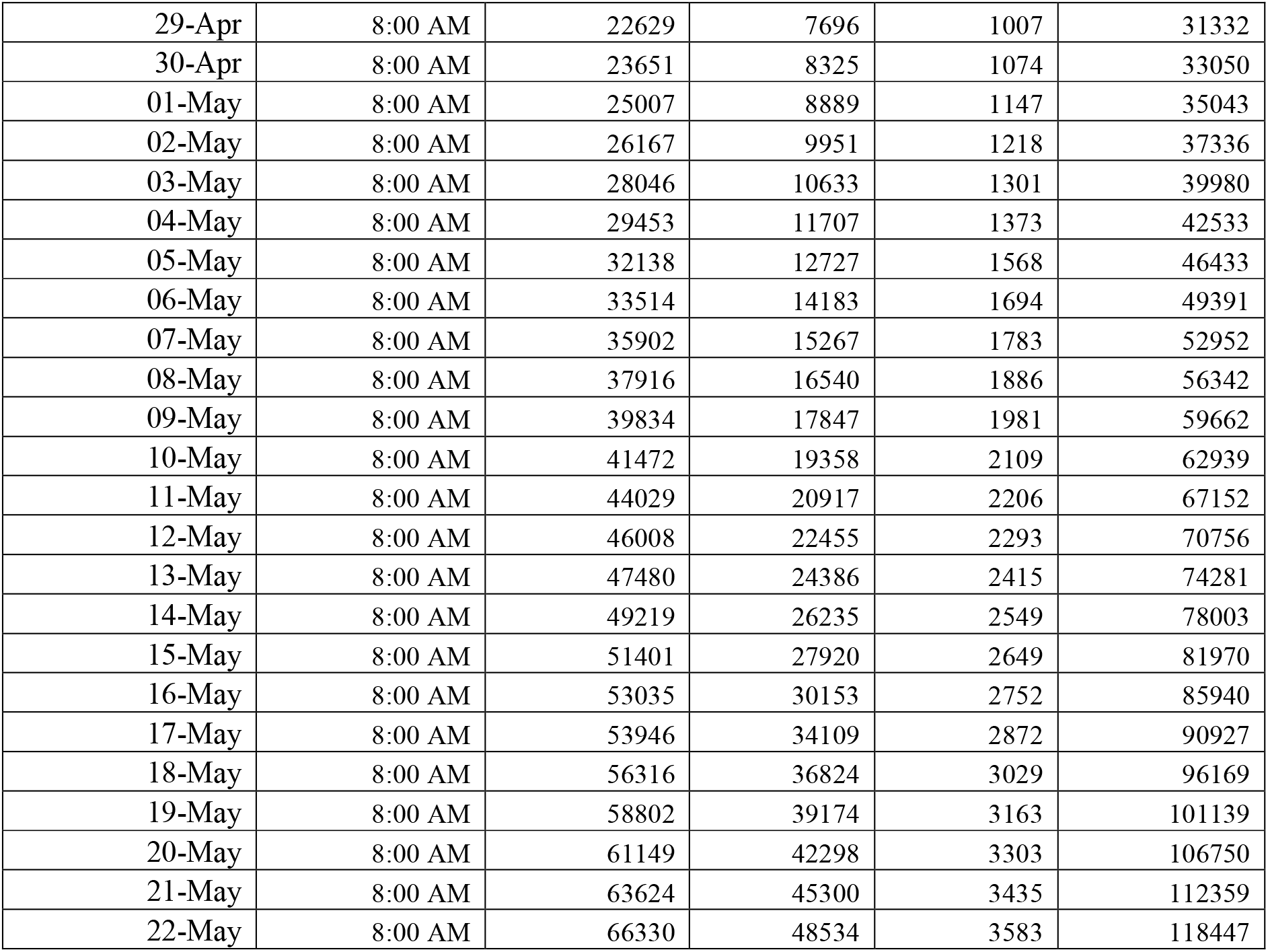
COVID-19 Cases in India

| Date | Time | Active cases | Total cured | Total death | Total cases |
| --- | --- | --- | --- | --- | --- |
| 01-Mar | 8:45 AM | 3 | 0 | 0 | 3 |
| 02-Mar | 8:45 AM | 3 | 0 | 0 | 3 |
| 03-Mar | 8:45 AM | 4 | 0 | 0 | 4 |
| 04-Mar | 8:45 AM | 26 | 0 | 0 | 26 |
| 05-Mar | 8:45 AM | 27 | 0 | 0 | 27 |
| 06-Mar | 8:45 AM | 28 | 0 | 0 | 28 |
| 07-Mar | 8:45 AM | 31 | 0 | 0 | 31 |
| 08-Mar | 8:45 AM | 37 | 0 | 0 | 37 |
| 09-Mar | 8:45 AM | 43 | 0 | 0 | 43 |
| 10-Mar | 8:45 AM | 58 | 0 | 0 | 58 |
| 11-Mar | 8:45 AM | 58 | 0 | 0 | 58 |
| 12-Mar | 8:45 AM | 69 | 0 | 1 | 70 |
| 13-Mar | 8:45 AM | 70 | 6 | 2 | 78 |
| 14-Mar | 8:45 AM | 88 | 6 | 2 | 96 |
| 15-Mar | 8:45 AM | 99 | 9 | 2 | 110 |
| 16-Mar | 8:45 AM | 114 | 9 | 2 | 125 |
| 17-Mar | 8:45 AM | 126 | 10 | 3 | 139 |
| 18-Mar | 8:45 AM | 152 | 10 | 3 | 165 |
| 19-Mar | 8:45 AM | 170 | 16 | 4 | 190 |
| 20-Mar | 8:45 AM | 221 | 19 | 5 | 245 |
| 21-Mar | 8:45 AM | 282 | 19 | 5 | 306 |
| 22-Mar | 8:45 AM | 304 | 20 | 7 | 331 |
| 23-Mar | 8:45 AM | 360 | 30 | 9 | 399 |
| 24-Mar | 8:45 AM | 446 | 37 | 9 | 492 |
| 25-Mar | 9:15 AM | 512 | 40 | 9 | 561 |
| 26-Mar | 10:15 AM | 593 | 42 | 13 | 648 |
| 27-Mar | 9:15 AM | 640 | 67 | 17 | 724 |
| 28-Mar | 9:30 AM | 775 | 79 | 19 | 873 |
| 29-Mar | 10:00 AM | 867 | 87 | 25 | 979 |
| 30-Mar | 10:30 AM | 942 | 100 | 29 | 1071 |
| 31-Mar | 8:30 PM | 1238 | 124 | 35 | 1397 |
| 01-Apr | 9:00 AM | 1466 | 133 | 38 | 1637 |
| 02-Apr | 9:00 AM | 1764 | 151 | 50 | 1965 |
| 03-Apr | 9:00 AM | 2088 | 157 | 56 | 2301 |
| 04-Apr | 9:00 AM | 2650 | 184 | 68 | 2902 |
| 05-Apr | 9:00 AM | 3030 | 267 | 77 | 3374 |
| 06-Apr | 9:00 AM | 3666 | 292 | 109 | 4067 |
| 07-Apr | 9:00 AM | 3981 | 326 | 114 | 4421 |
| 08-Apr | 9:00 AM | 4643 | 402 | 149 | 5194 |
| 09-Apr | 8:00 AM | 5095 | 473 | 166 | 5734 |
| 10-Apr | 8:00 AM | 5709 | 504 | 199 | 6412 |
| 11-Apr | 8:00 AM | 6565 | 643 | 239 | 7447 |
| 12-Apr | 8:00 AM | 7367 | 716 | 273 | 8356 |
| 13-Apr | 8:00 AM | 7987 | 857 | 308 | 9152 |
| 14-Apr | 8:00 AM | 8988 | 1036 | 339 | 10531 |
| 15-Apr | 8:00 AM | 9756 | 1306 | 377 | 11439 |
| 16-Apr | 8:00 AM | 10477 | 1489 | 414 | 12380 |
| 17-Apr | 8:00 AM | 11201 | 1749 | 437 | 13387 |
| 18-Apr | 8:00 AM | 11906 | 1992 | 480 | 14378 |
| 19-Apr | 8:00 AM | 12974 | 2231 | 507 | 15712 |
| 20-Apr | 8:00 AM | 14175 | 2547 | 543 | 17265 |
| 21-Apr | 8:00 AM | 14759 | 3252 | 590 | 18601 |
| 22-Apr | 8:00 AM | 15474 | 3870 | 640 | 19984 |
| 23-Apr | 8:00 AM | 16454 | 4258 | 681 | 21393 |
| 24-Apr | 8:00 AM | 17610 | 4749 | 718 | 23077 |
| 25-Apr | 8:00 AM | 18668 | 5063 | 775 | 24506 |
| 26-Apr | 8:00 AM | 19868 | 5804 | 824 | 26496 |
| 27-Apr | 8:00 AM | 20835 | 6185 | 872 | 27892 |
| 28-Apr | 8:00 AM | 21632 | 6869 | 934 | 29435 |
| 29-Apr | 8:00 AM | 22629 | 7696 | 1007 | 31332 |
| 30-Apr | 8:00 AM | 23651 | 8325 | 1074 | 33050 |
| 01-May | 8:00 AM | 25007 | 8889 | 1147 | 35043 |
| 02-May | 8:00 AM | 26167 | 9951 | 1218 | 37336 |
| 03-May | 8:00 AM | 28046 | 10633 | 1301 | 39980 |
| 04-May | 8:00 AM | 29453 | 11707 | 1373 | 42533 |
| 05-May | 8:00 AM | 32138 | 12727 | 1568 | 46433 |
| 06-May | 8:00 AM | 33514 | 14183 | 1694 | 49391 |
| 07-May | 8:00 AM | 35902 | 15267 | 1783 | 52952 |
| 08-May | 8:00 AM | 37916 | 16540 | 1886 | 56342 |
| 09-May | 8:00 AM | 39834 | 17847 | 1981 | 59662 |
| 10-May | 8:00 AM | 41472 | 19358 | 2109 | 62939 |
| 11-May | 8:00 AM | 44029 | 20917 | 2206 | 67152 |
| 12-May | 8:00 AM | 46008 | 22455 | 2293 | 70756 |
| 13-May | 8:00 AM | 47480 | 24386 | 2415 | 74281 |
| 14-May | 8:00 AM | 49219 | 26235 | 2549 | 78003 |
| 15-May | 8:00 AM | 51401 | 27920 | 2649 | 81970 |
| 16-May | 8:00 AM | 53035 | 30153 | 2752 | 85940 |
| 17-May | 8:00 AM | 53946 | 34109 | 2872 | 90927 |
| 18-May | 8:00 AM | 56316 | 36824 | 3029 | 96169 |
| 19-May | 8:00 AM | 58802 | 39174 | 3163 | 101139 |
| 20-May | 8:00 AM | 61149 | 42298 | 3303 | 106750 |
| 21-May | 8:00 AM | 63624 | 45300 | 3435 | 112359 |
| 22-May | 8:00 AM | 66330 | 48534 | 3583 | 118447 |

State wise COVID-19 disease cases are shown in Fig. 2 [18]. It can be analyzed from the Fig. 2 that the Maharashtra, Gujarat, Delhi, Tamil Nadu, Telangana, Rajasthan, Uttar Pradesh, Madhya Pradesh, Kerala and Andhra Pradesh are most suffered state in the country

**Figure 2:**
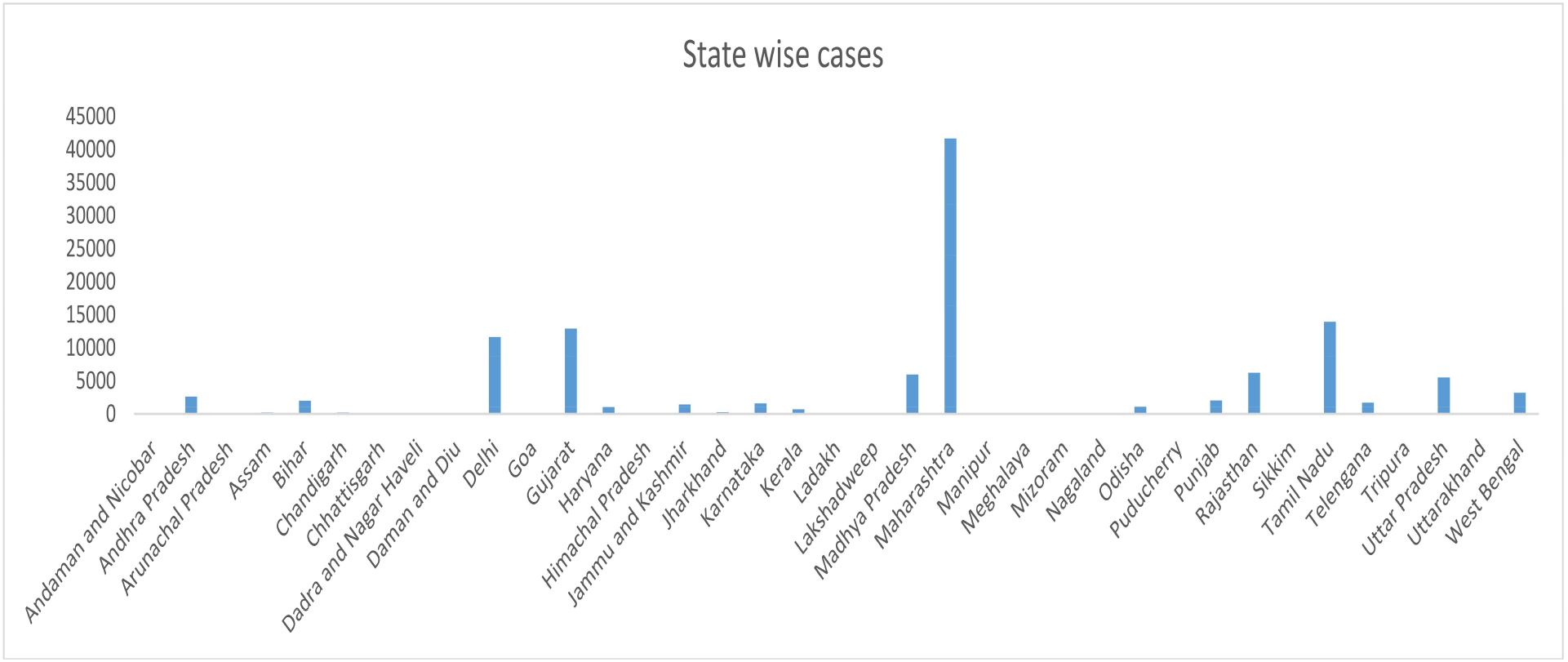
State wise COVID-19 cases in India

## 4. Result and Discussion

Initially, the natural evolution of the pandemic impacting the Indian population was modelled, without catering for any NPI or other interventions. For ease of modelling, we have assumed homogenous distribution of the Indian population that does not capture variations in population density or the urban-rural variations. It is also being assumed that all the peoples have equal susceptibility to COVID-19.

The mathematical model 1 has been modelled to implement natural evolution of the COVID-19 disease in the Indian population was without screening, quarantine and NPI. It has been reported in literature that multiplying factor for COVID-19 is approximately 2.6. Hence, the spread scenario has been shown in Fig. 3 and Fig. 4 for different number of days. The growth of COVID-19 cases is exponential in the case of without screening, quarantine and NPI.

**Figure 3:**
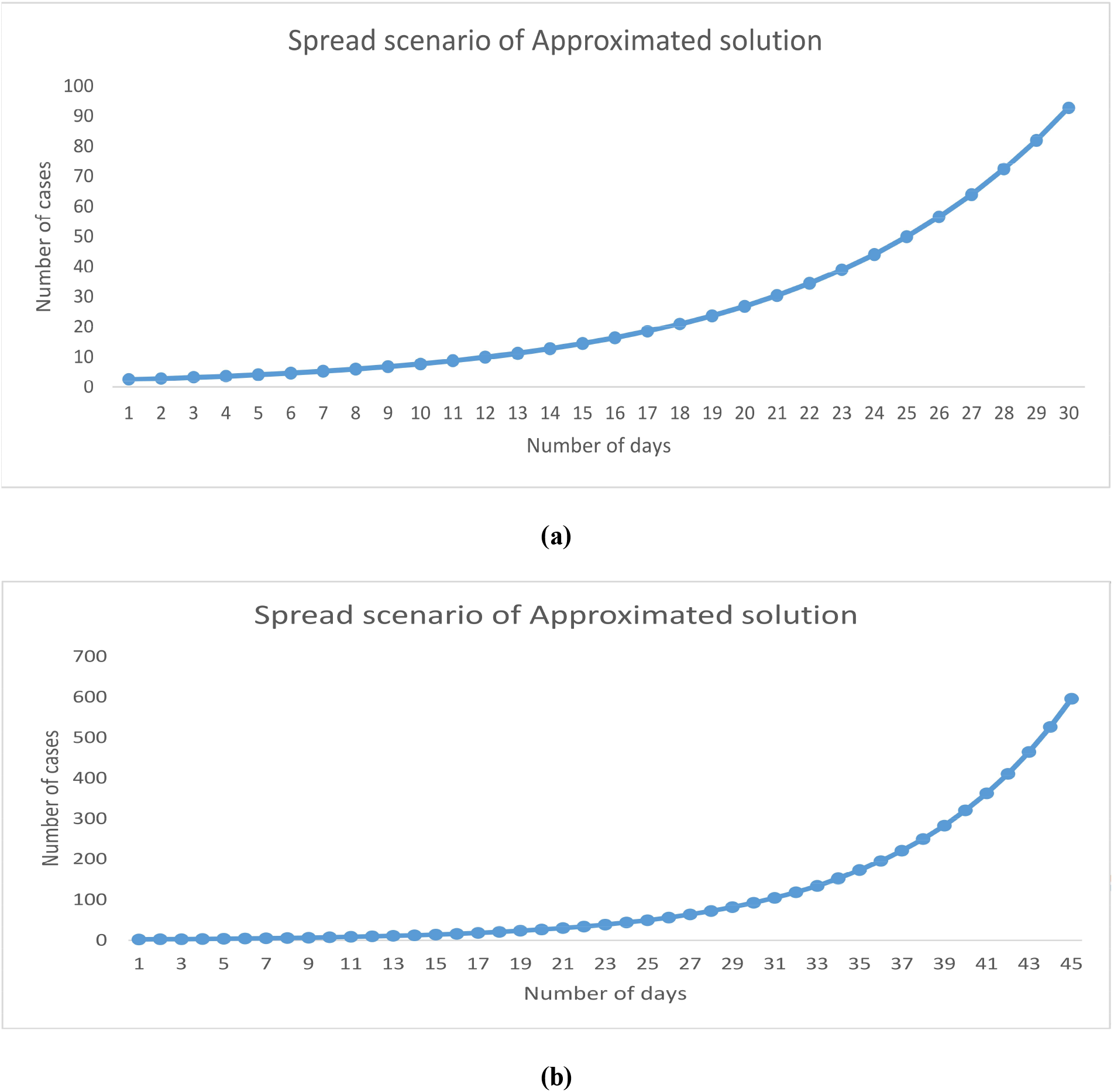
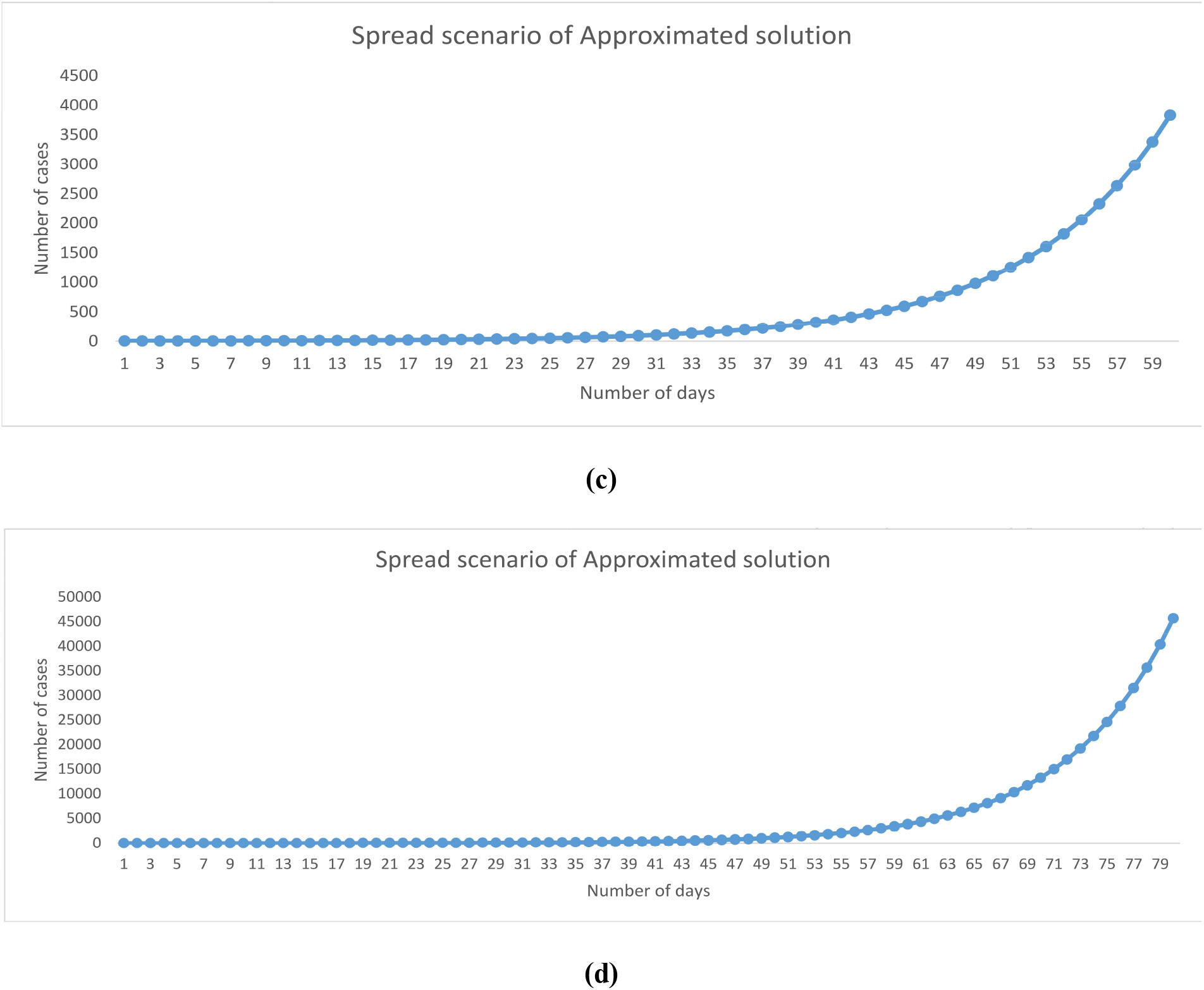
Covid-19 spread scenario without screening, quarantine and Lockdown for (a) 30 days (b) 45 days (c) 60 days (d) 80 days

**Figure 4:**
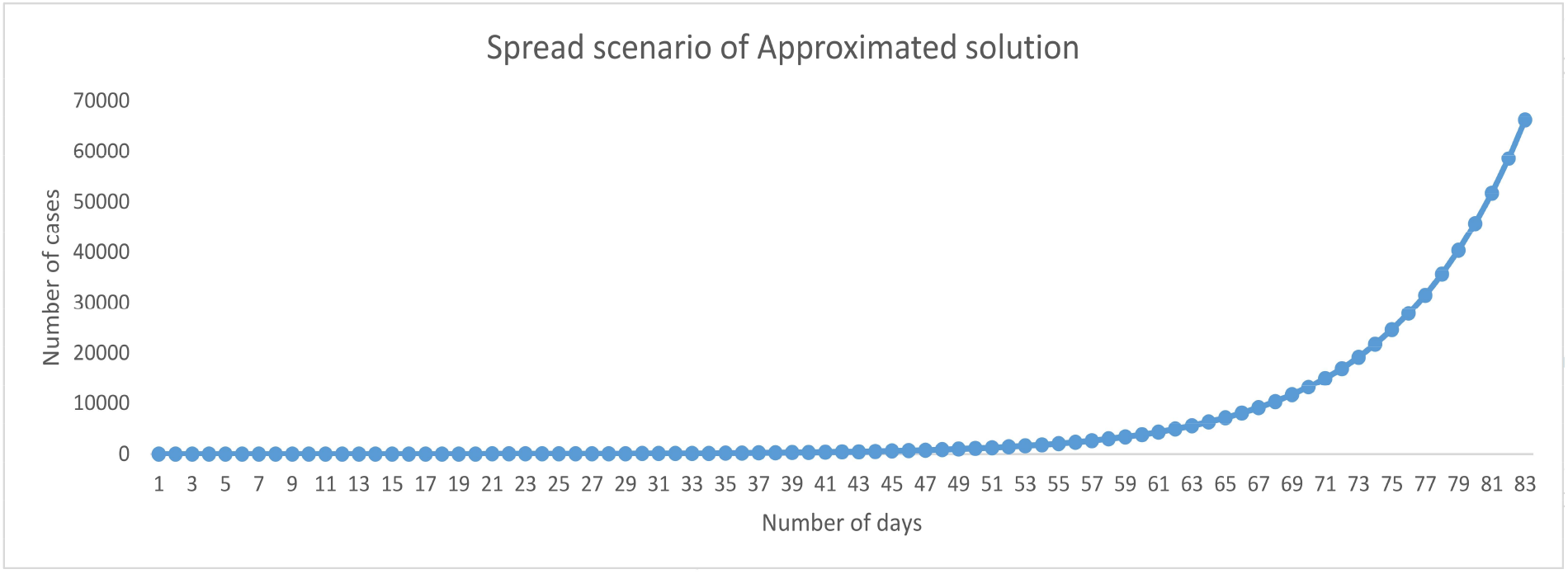
Covid-19 spread scenario without screening, quarantine and Lockdown for 83 days

Screening and Quarantine measures are followed to reduce spread. India has started the screening as early as in early of January 2020. All the travelers coming from selected countries have been screened at international airport.

These travelers have been further divided in quarantine group and normal group dep ending on the condition at time of inspection. This condition represented by proposed model 2. At this stage, NPI interventions were not there or present in minimal amount. It can be noticed from model 2 that the total number of cases is summation of quarantine cases and leakage. It can also be noticed that leakage that exponential behavior. The multiplication term present in leakage reduces the growth of the function. It can also be noticed from the Fig.5 and Fig. 6. Fig. 5 and Fig. 6 are representing the spread scenario before NPI interventions. Only screening and quarantine were in place. The growth shows a random behavior due to small contribution from leakage factor. The quarantine cases follow a linear nature and have large impact on total cases and active cases initially. Impact of leakage factor increases with time and the graph is also showing similar behavior. Table 1 presents the data for India. It has been plotted to have visual results. Fig. 7 shows the total number of infected persons who have been cured from this disease. It can be noticed that plot is showing a growth with time and more people are coming out from this pandemic. Fig. 8 represents total death occurred. It can also be noticed that with the time number of death are also increasing. It has been found that the growth of death curve is lesser than the growth of total cured. Fig. 9 and Fig. 10 present the active cases and total cases of COVID-19 in India respectively. Curve fitting has been used to obtain the best fitted equation for the data. It has been reported that exponential curve fitting is providing go o d results with approximately 96% accuracy. Although, active cases and total cases is following exponential behavior, yet the growth is in the order of 0.17 approximately for active cases as well as for total cases also. This shows that the measures taken by Indian government has slow down the growth of COVID-19 spread. The NPI measures have been imposed on the Indian population from March 24, 2020. Hence, impact of these NPI measures are also of interest for research. This impact has been studied in current manuscript. The mathematical model 3 is following screening, quarantine and NPI. Quarantine and NPI measure are being implemented to reduce spreading. The numbers of freshly infected decreases with the increase in quarantined cases in general. The effectiveness of quarantine dep ends on the compliance by individuals and public health measures that are instituted. The results obtained by this model are shown in the table 2. It shows the number of active cases and total cases after imposing NPI measures. Fig. 11 and Fig. 12 represent the active cases and total cases after NPI measures such as Lockdown, social distancing etc. It can be analyzed for the Fig. 11 and Fig. 12 that lockdown reduces spread exotically.

**Figure 5:**
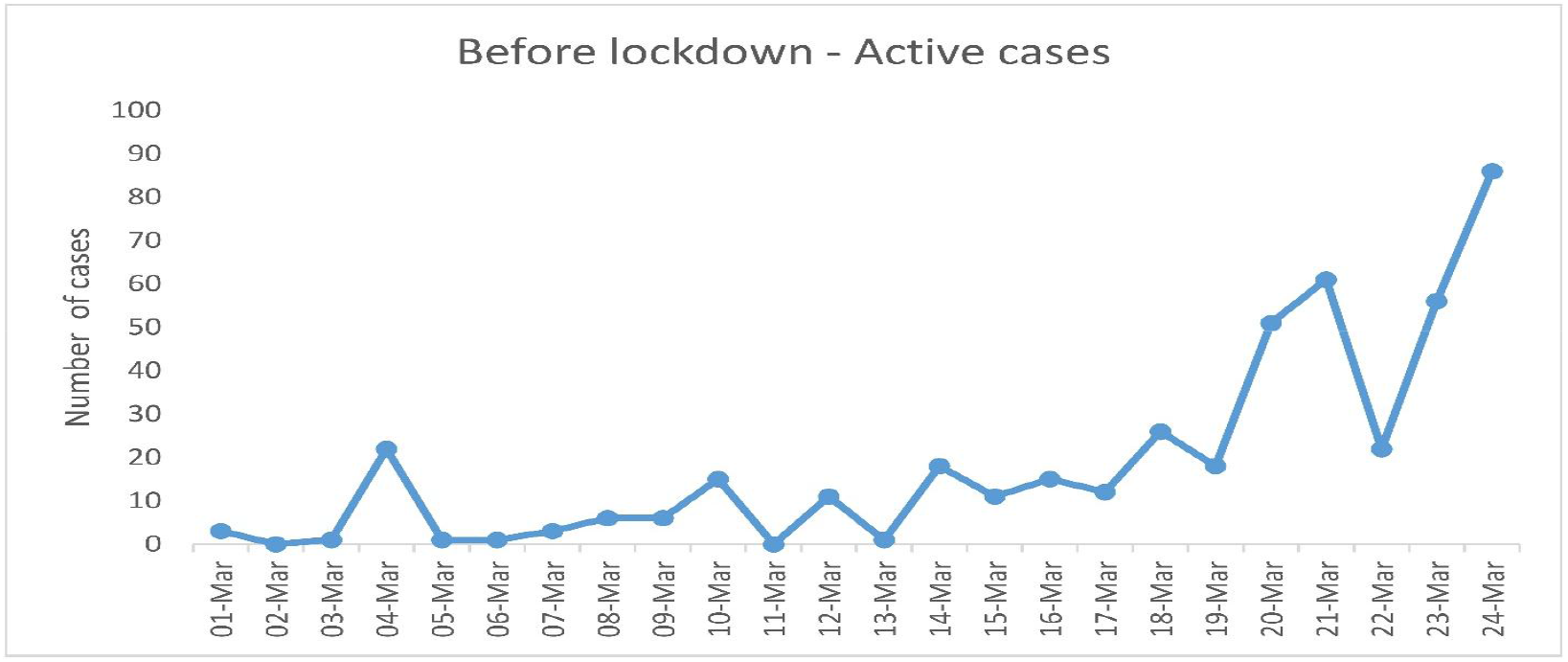
COVID-19 Active Cases Spread Scenario for India before Lockdown

**Figure 6:**
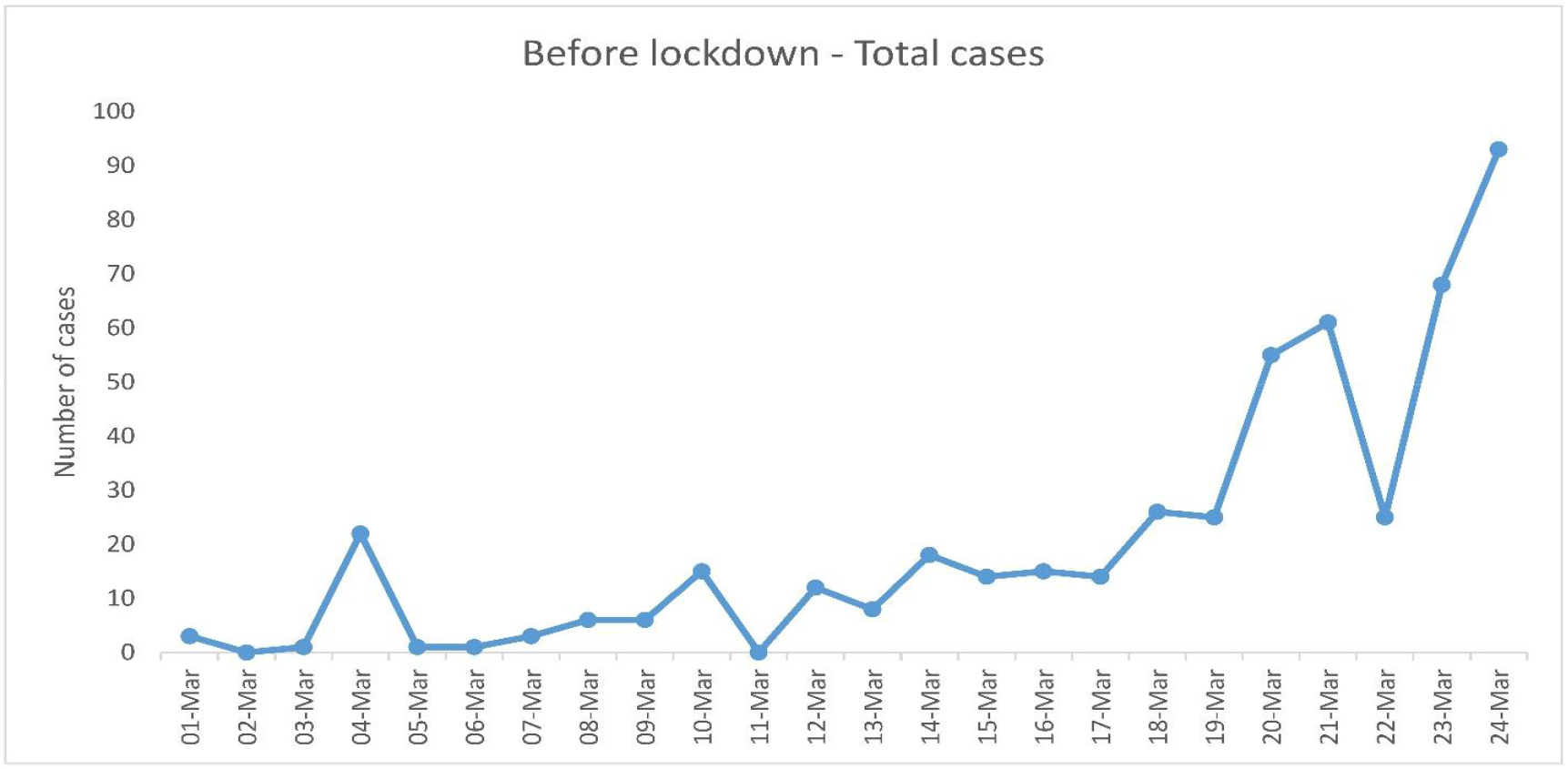
COVID-19 Total Cases Spread Scenario for India before Lockdown

**Figure 7:**
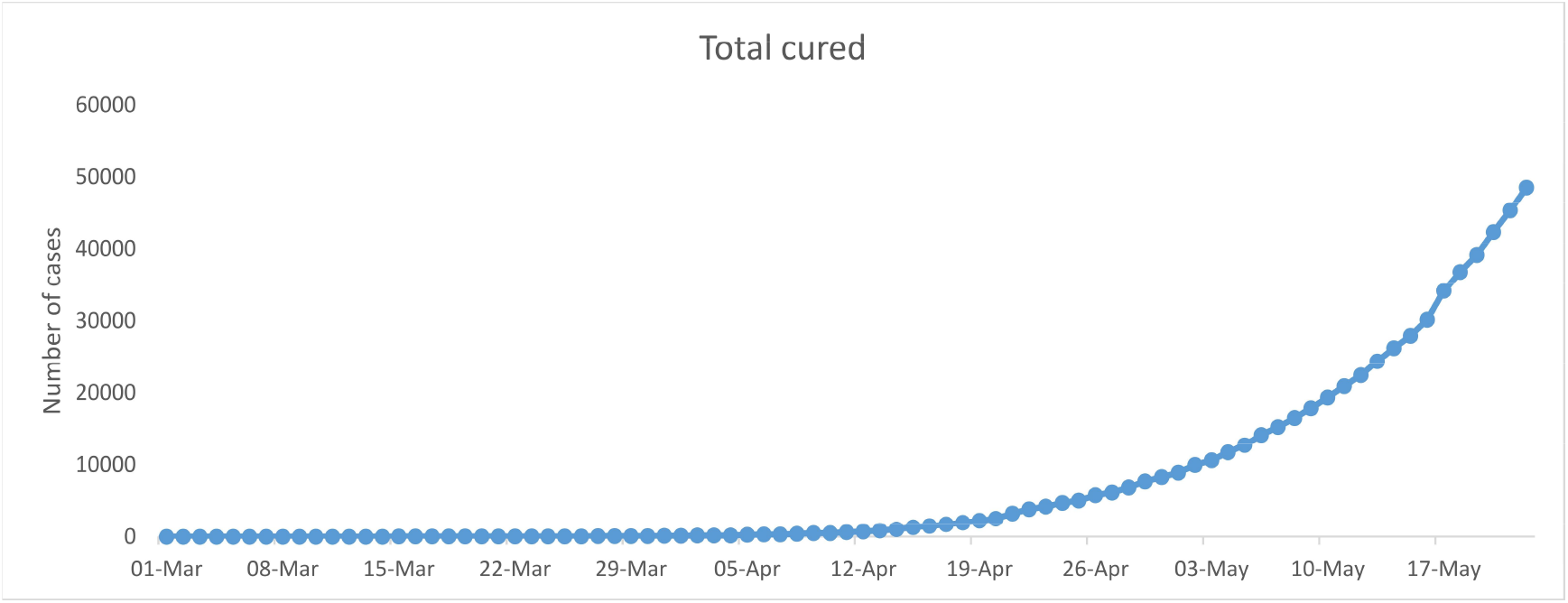
Total COVID-19 cured cases

**Figure 8:**
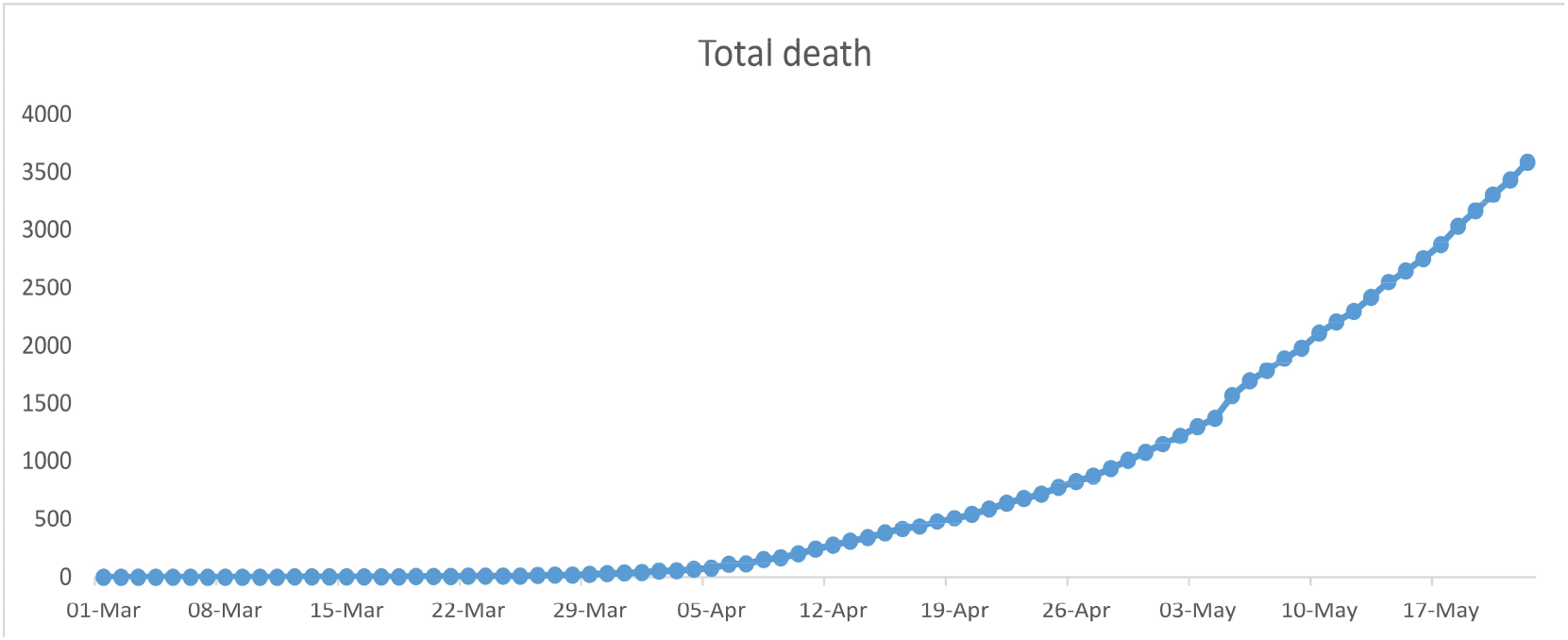
Total COVID-19 death cases

**Figure 9:**
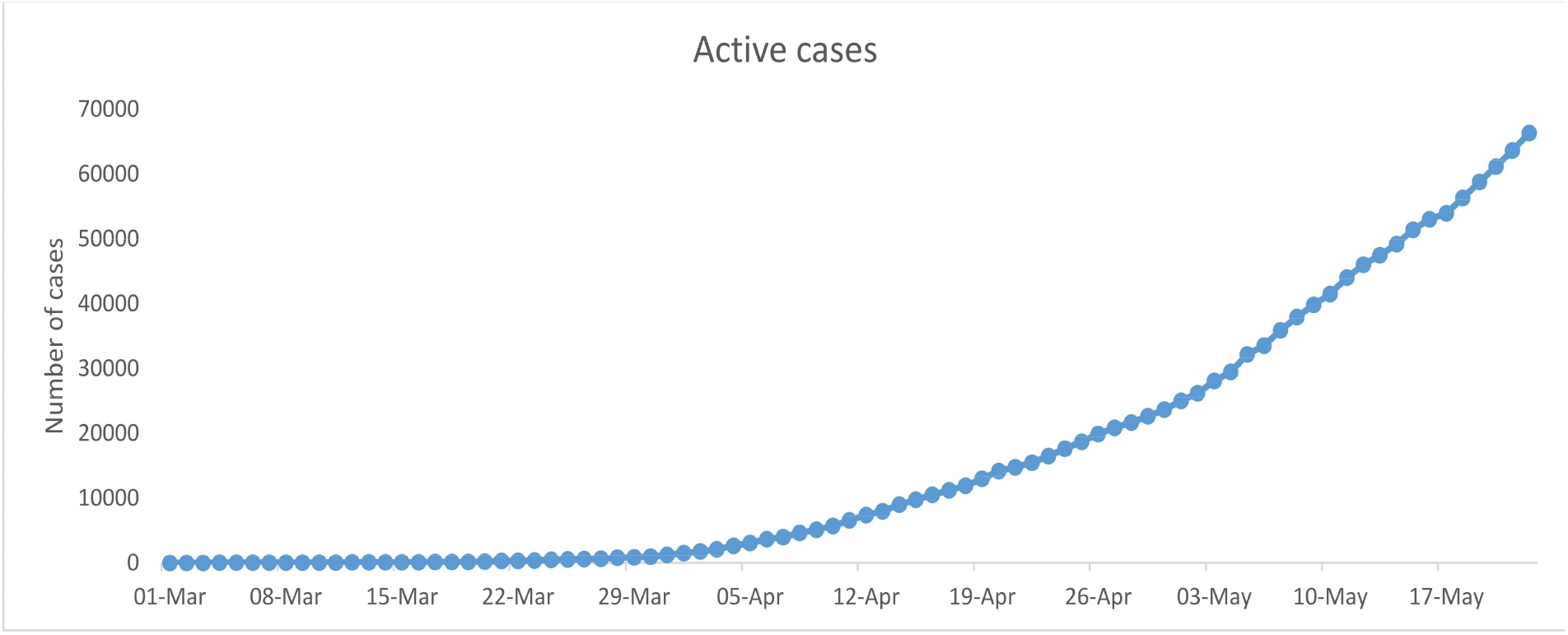
Active COVID-19 cases

**Figure 10:**
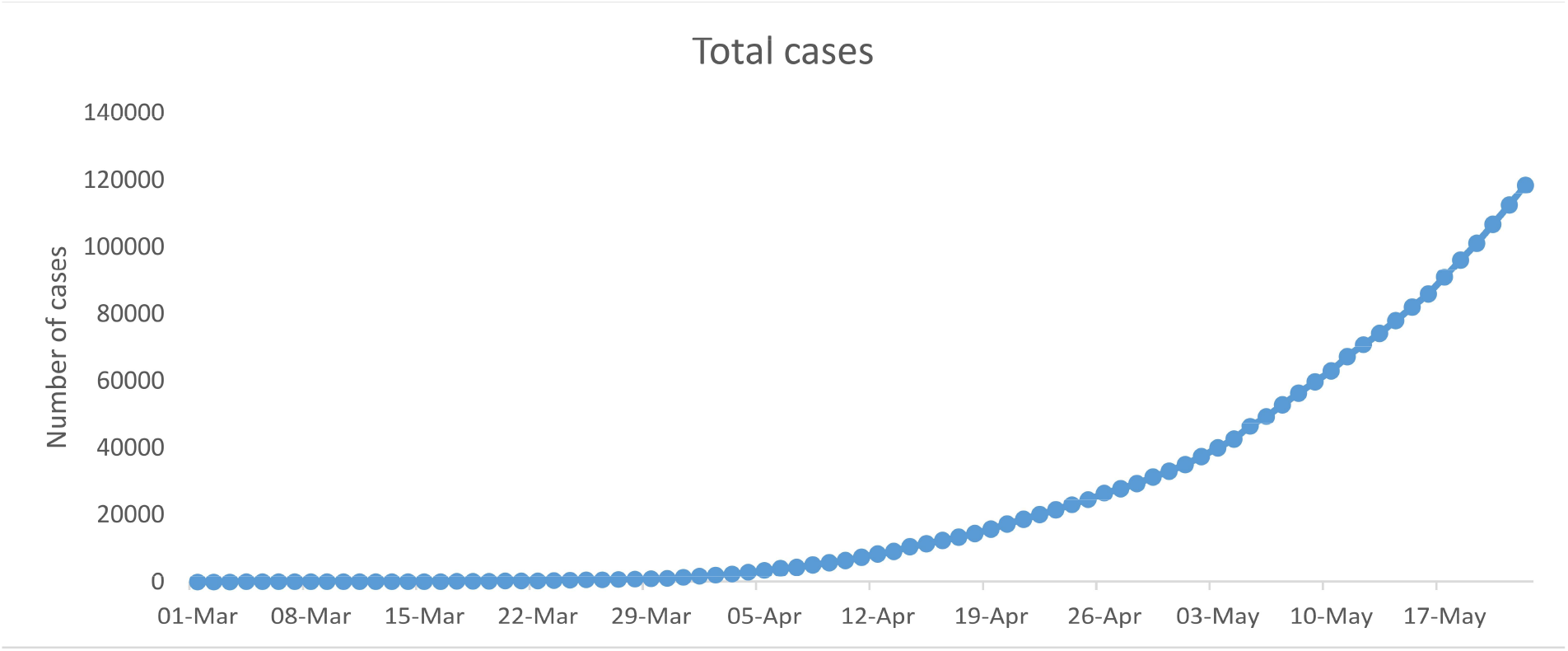
Total Cases of COVID-19

**Table 2:**
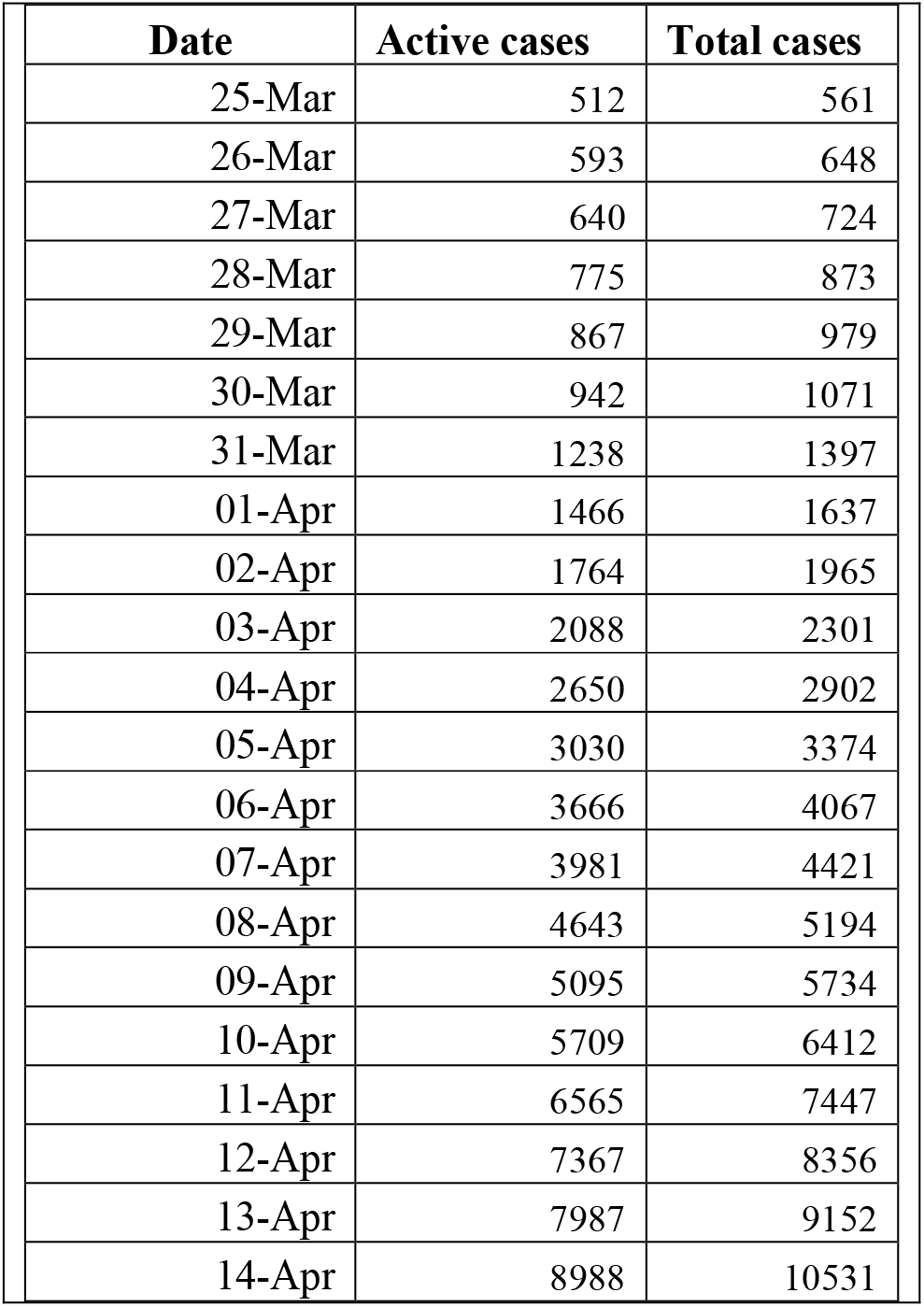
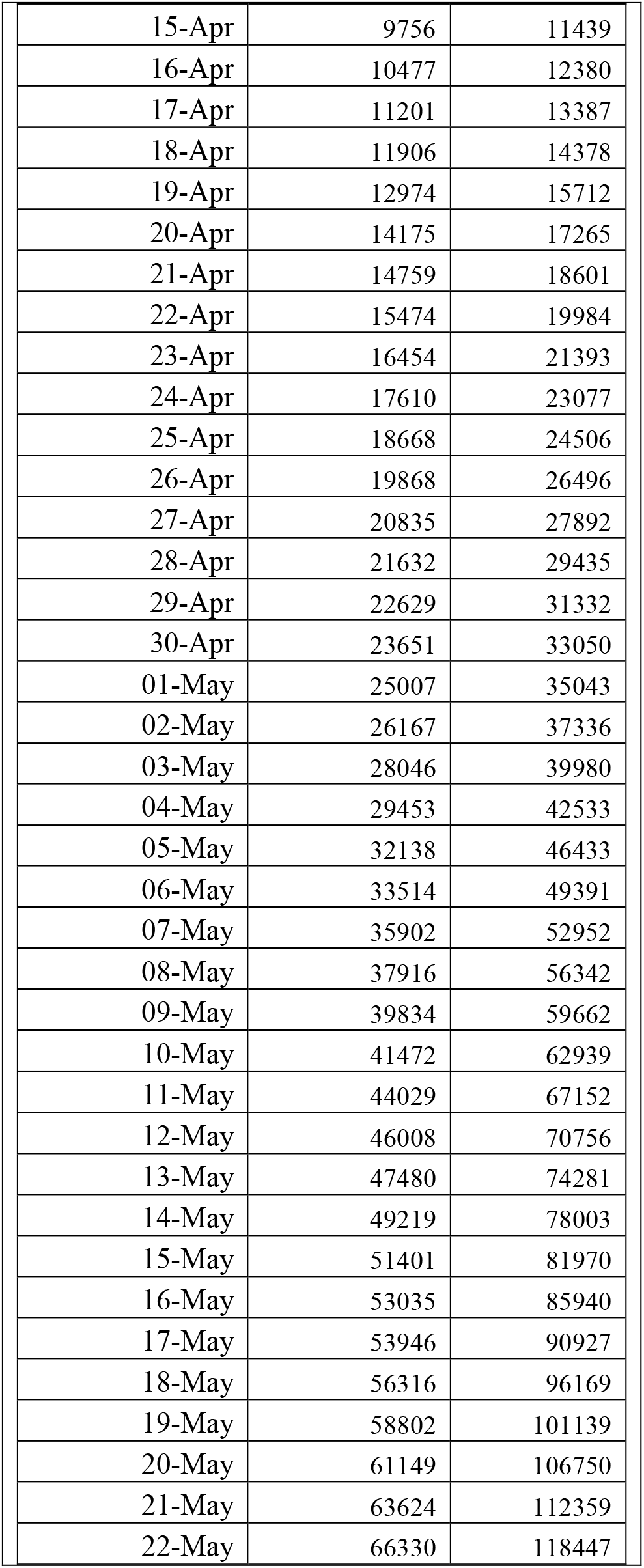
Active cases and Total cases after lockdown

**Figure 11:**
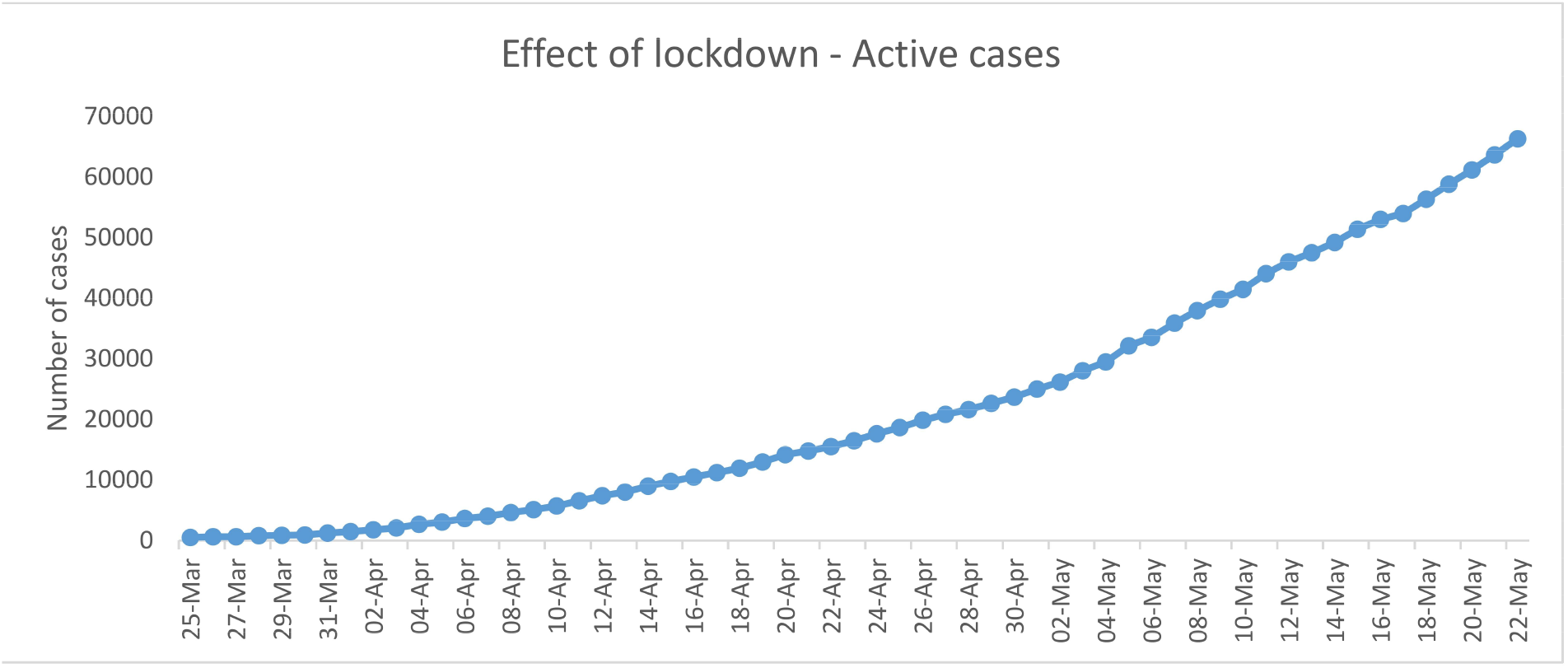
Effect on Active Cases on COVID-19 Data Spread Scenario for India after lockdown

**Figure 12:**
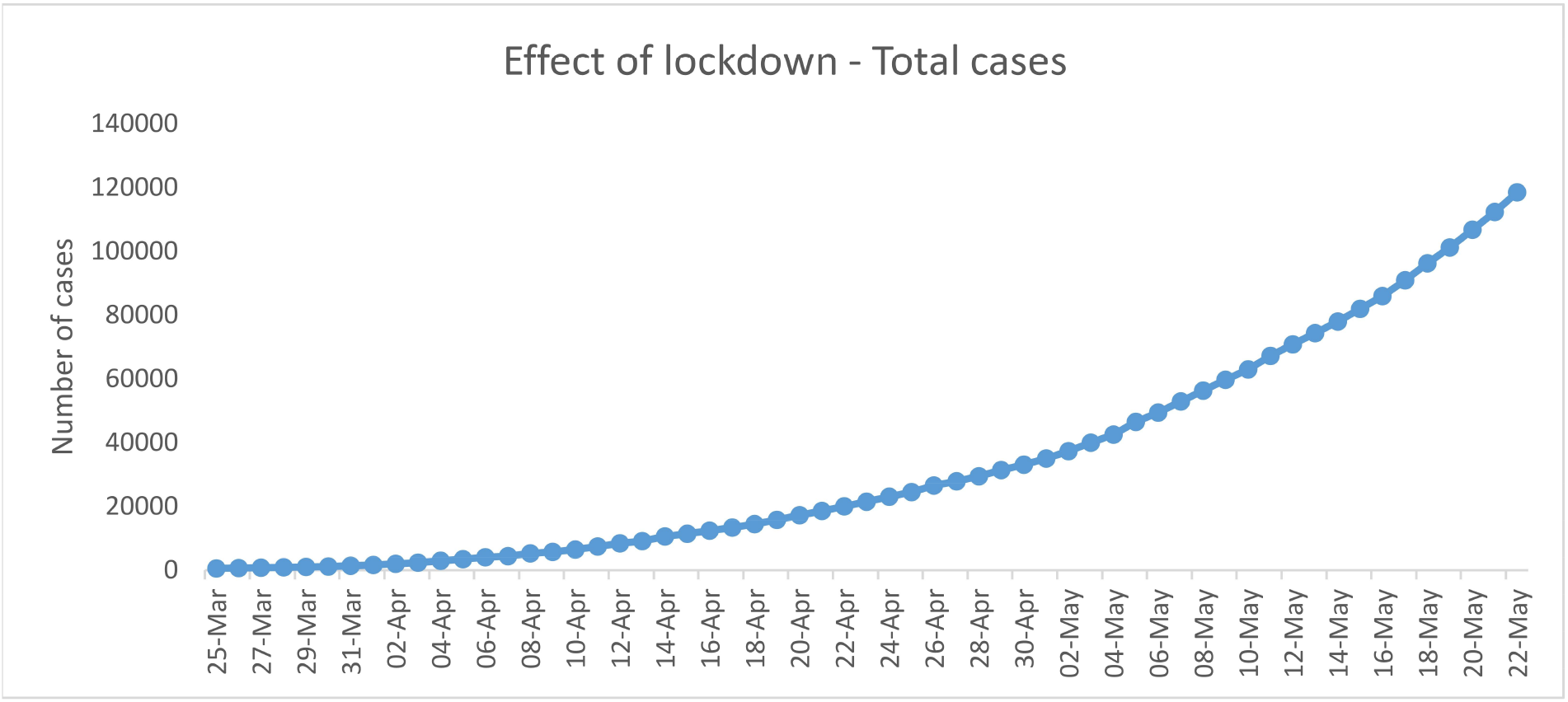
Effect on Total Cases on COVID-19 Data Spread Scenario for India After lockdown

Table 3 and 4 presents the R2 values for different curve fitting methods applied on Fig. 11 and Fig. 12 for active cases and total cases respectively. T has been found that polynomial fitting of order 2 is providing good fitting results for both active and total cases scenario. This indicates that the growth of active and total cases reduces from exponential to polynomial.

**Table 3:** Active cases after lockdown

| Types of curve | Equation | R-squared value(R<sup>2</sup>) |
| --- | --- | --- |
| Exponential function | $y = 1086e^{0.0788x}$ | 0.9297 |
| Linear function | $y = 1112.4x - 11111$ | 0.9284 |
| Logarithmic function | $y = 17228\ln(x) - 31623$ | 0.6111 |
| Polynomial of order 2 | $y = 20.098x^2 - 93.452x + 1148.4$ | 0.9987 |
| Polynomial of order 3 | $y = 0.0922x^3 + 11.802x^2 + 107.32x + 102.62$ | 0.999 |
| Polynomial of order 4 | $y = -0.0018x^4 + 0.3091x^3 + 3.3883x^2 + 221.74x - 266.53$ | 0.999 |

**Table 4:** Total cases after lockdown

| Types of curve | Equation | R-squared value(R<sup>2</sup>) |
| --- | --- | --- |
| Exponential function | $y = 1099.4e^{0.0875x}$ | 0.9517 |
| Linear function | $y = 1847.5x - 21754$ | 0.8753 |
| Logarithmic function | $y = 27736\ln(x) - 53078$ | 0.5414 |
| Polynomial of order 2 | $y = 44.898x^2 - 846.35x + 5633.3$ | 0.9952 |
| Polynomial of order 3 | $y = 0.5811x^3 - 7.3996x^2 + 419.35x - 959.62$ | 0.9996 |
| Polynomial of order 4 | $y = 0.0063x^4 - 0.1736x^3 + 21.877x^2 + 21.24x + 324.83$ | 0.9998 |

Hence, NPI measures has reduces or slowdown the growth of COVID-19 which has been also indicated in Fig 6. Complete lockdown represents full implementation of NPI measures to entire population. Our model shows that immediate implementation of effective combination of screening, quarantine and NPIs reduces COVID-19 disease drastically. Hence, the numbers infected peoples with COVID-19 become manageable, reducing from millions to merely thousands.

The overall COVID-19 spread in India has been shown in Fig. 13 along with approximation obtained by proposed method. The lockdown period required by the India has been analyzed as per growth rate obtained. It has been shown in the Fig. 14. It can be analyzed from the Fig. 14 that there will be no COVID-19 cases after 13-July-2020.

**Figure 13:**
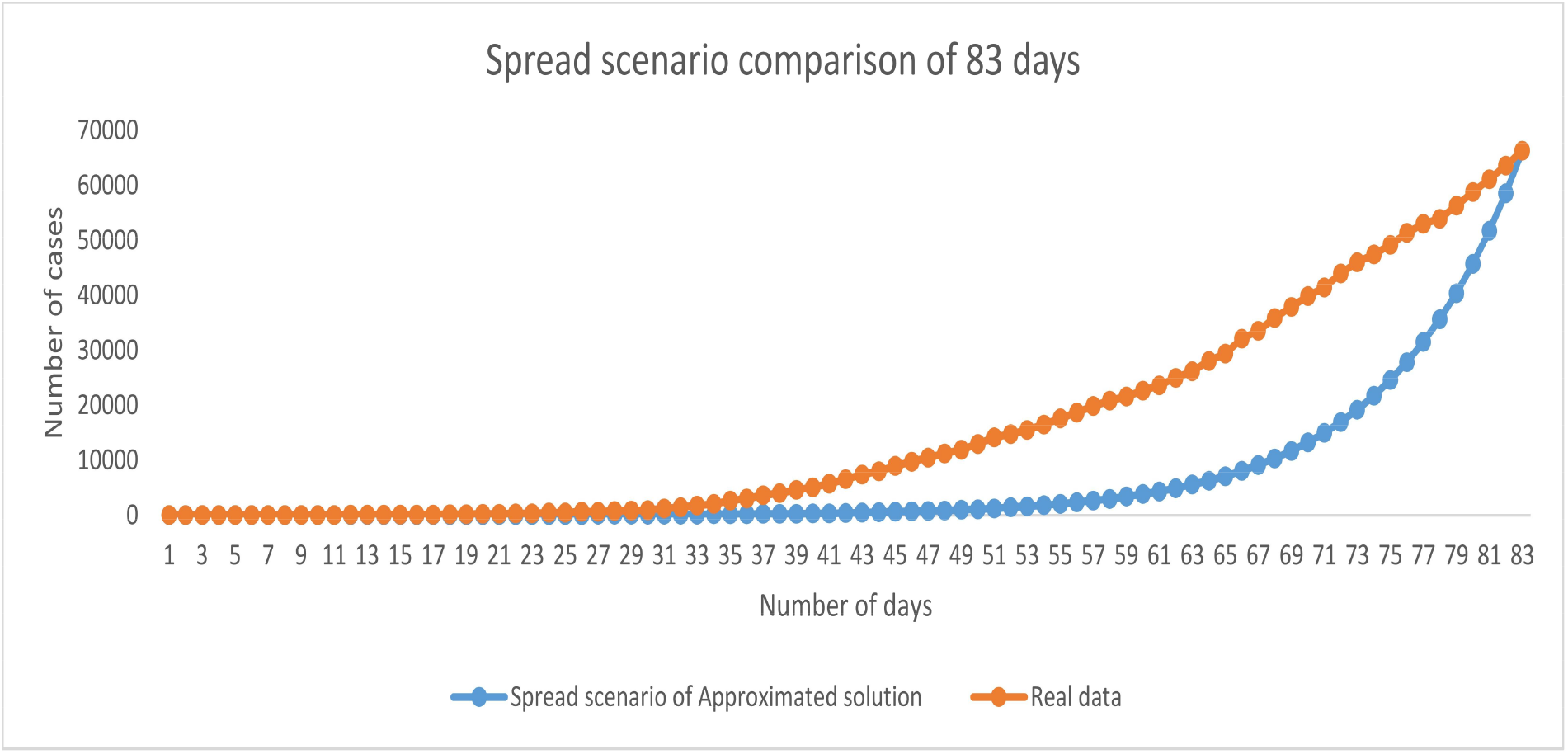
COVID-19 spread approximation by proposed method

**Figure 14:**
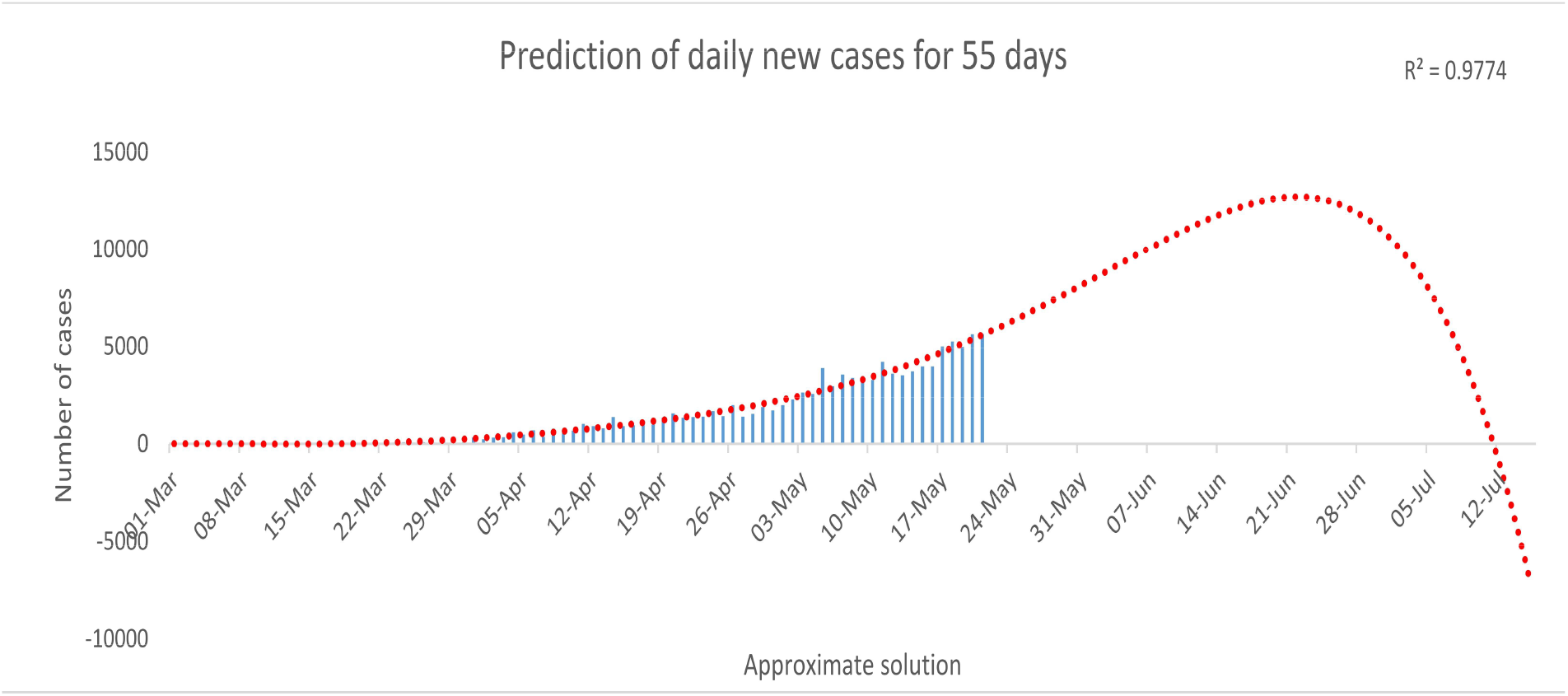
Total lockdown period required as per growth rate

## 5 Conclusion

India is in the early stage of the COVID-19 pandemic, with a lower growth rate than other countries studied. In the present manuscript, three mathematical model have been presented to estimate growth function of COVID-19 disease. It has been found that the numbers of COVID-19 patients will be more without screening the peoples coming from other countries. Since, every people suffering with COVID-19 disease are spreaders. The screening and quarantine have been implemented in mathematical model 2. It has been found that number of spreaders is less than that of model 1. However, expected performance is not achieved due to leakage i.e. peoples not following quarantine. Mathematical model 3 has been designed with screening, quarantine with NPIs. It has achieved reasonable performance in reducing number of spreaders. Hence, it is found that immediate implementation of Non-Pharmacological Interventions among the general population, including complete lockdowns, has the potential to retard the progress of the pandemic. It has been also observed that COVID-19 spread will be recovered till 13-July-2020 approximately in India.

## Data Availability

Indian government websites:
https://www.mohfw.gov.in

## Acknowledgment

The work has been supported by a grant received from the Ministry of Education, Government of India under the Scheme for the Promotion of Academic and Research Collaboration (SPARC), 2019.

